# An Artificial Intelligence-Assisted Diagnostic Platform for Rapid Near-Patient Hematology

**DOI:** 10.1101/2021.04.27.21255770

**Authors:** Neta Bachar, Dana Benbassat, David Brailovsky, Yochay Eshel, Dan Glück, Daniel Levner, Sarah Levy, Sharon Pecker, Evgeny Yurkovsky, Amir Zait, Cordelia Sever, Alexander Kratz, Carlo Brugnara

**Affiliations:** S. D. Sight Diagnostics LTD, 23 Begin Menachem Road, Tel Aviv-Jaffa, Israel; TriCore Reference Laboratories, Albuquerque, NM, USA; Department of Pathology and Cell Biology, Columbia University College of Physicians and Surgeons NY, New York, USA; New York-Presbyterian Hospital, NY, New York, USA; Department of Laboratory Medicine, Boston Children’s Hospital and Department of Pathology, Harvard Medical School, Boston, MA, USA

## Abstract

Hematology analyzers capable of performing complete blood count (CBC) have lagged in their prevalence at the point-of-care. Sight OLO® (Sight Diagnostics, Israel) is a novel hematological platform which provides a 19 parameter, five-part differential CBC, and is designed to address the limitations in current point-of-care hematology analyzers using recent advances in artificial intelligence (AI) and computer vision. Accuracy, repeatability, and flagging capabilities of OLO were compared with the Sysmex XN-Series System (Sysmex, Japan). Matrix studies compared performance using venous, capillary and direct-from-finger-prick blood samples. Regression analysis shows strong concordance between OLO and the Sysmex XN, demonstrating that OLO performs with high accuracy for all CBC parameters. High repeatability and reproducibility were demonstrated for most of the testing parameters. The analytical performance of the OLO hematology analyzer was validated in a multicenter clinical laboratory setting, demonstrating its accuracy and comparability to clinical laboratory-based hematology analyzers. Furthermore, the study demonstrated the validity of CBC analysis of samples collected directly from fingerpricks.

**One Sentence Summary:** We present a novel diagnostic platform based on artificial intelligence-assisted image analysis that is capable of performing rapid complete blood count from venous, capillary, and finger-prick samples in near-patient settings.

## INTRODUCTION

A large number of laboratory tests are now available for point-of-care testing (POCT)(1) including, among others, urinalysis, blood chemistry and infectious disease testing (2–5). However, hematology analyzers capable of performing complete blood counts (CBCs) have lagged in their prevalence at the point-of-care(6), despite CBCs ranking among the most commonly ordered tests. The relative absence of POC CBC analyzers has also hampered the adoption of other POC tests that tend to be ordered alongside the CBC (e.g. complete metabolic panels); in particular, if a CBC is required but must be performed in the central lab due to its absence at the point-of-care, its companion tests may as well be ordered from the central lab.

The majority of current near-patient CBC analyzers rely on the miniaturization of large-lab analyzers, particularly those based on flow cytometry and/or impedance cytometry techniques(1, 4, 7). Such miniaturization has in many cases been accompanied by simplification, which has resulted in reduced numbers of parameters (e.g. three-part instead of five-part differentials), narrower reportable ranges, reduced abnormal cell flagging capabilities and in some cases a reduction in accuracy (as indicated by the small number of FDA clearances)(8–10). These CBC analyzers have also inherited many of the product attributes of their larger relatives, including the requirements for liquid reagent replacement, washout and calibration procedures, and frequent quality-control processes(11)which in the POC context can be limiting or prohibitive.

Sight OLO^®^ (S.D. Sight Diagnostics LTD, Israel) is a novel hematological platform designed to address some of the limitations in current near-patient and POCT hematology analyzers. OLO is based on recent advances in artificial intelligence (AI) and computerized image analysis (computer vision), and it provides a 19 parameter, five-part differential CBC. OLO employs single-use test-kits, a “dry” instrument and an extensive “Failsafe” self-test system to reduce operation overheads and simplify operation. OLO has received FDA 510(k) clearance for use in CLIA non-waived settings(12) and is CE Marked for POC use, including its use with samples collected directly from finger-pricks. Here, the results are presented for the accuracy of OLO as compared with the Sysmex XN-Series System (Sysmex, Kobe, Japan), as well as repeatability, flagging capabilities, reproducibility and matrix studies comparing the venous, capillary and finger-prick blood samples.

### The device

Sight OLO is a desktop hematology platform that was designed from the ground up for POC and near-patient settings. The device employs single-use test-kits that use a novel method for creating and staining blood smears within disposable test cartridges (please see “sample preparation” below). These monolayer blood smears are rapidly imaged using OLO’s automated fluorescence microscope to yield a set of more than 1,000 multispectral micrographs per sample. The images are analyzed using OLO’s onboard computer as they are collected using a set of specially designed algorithms, with the analysis resulting in the 19 reported CBC parameters and several flags, including flagging WBC for the presence of nucleated RBCs, blast cells, immature granulocytes and atypical lymphocytes. OLO then displays the diagnostic result on its touchscreen interface, optionally provides a paper printout and transmits the result digitally to a laboratory information management system (LIMS) or electronic medical records (EMR) system. The device measures 32 cm by 28 cm by 25 cm (about one cubic foot) and houses no reagents; reagents are contained within each test-kit, obviating the need for regular instrument washouts. OLO is factory calibrated so it requires no regular calibration. OLO’s Failsafe system works as an internal quality control to ensure that the device is performing appropriately, thereby obviating external quality control (QC) materials except as mandated by regulation.

### Sample requirements and preparation process

Sight OLO accepts both venous and finger-prick blood samples. In the finger-prick workflow, two drops of blood totaling 27µL are collected directly from the finger using the components provided in each single-use test-kit as described below. OLO was designed to work with low sample volumes to permit sample collection with no “milking” of the finger, in order to minimize patient discomfort. In the venous workflow (which also supports capillary samples collected in low-volume collection tubes), blood is provided in a K_2_EDTA tube; the sample is deposited from the collection tube onto hydrophobic paper using standard tube-top dispensers (e.g. Labcon U-Pette™) and then prepared using the components of the same test-kit and following the same process as finger-prick samples.

Each Sight OLO single-use test-kit contains a test cartridge, two capillaries for sample collection and a reagent-filled mixing-bottle. In turn, the test cartridge contains two separate sample chambers: a hemoglobin chamber and an imaging chamber. During sample preparation, a single drop of undiluted blood (17µL) is used to fill the hemoglobin chamber; in the finger-prick workflow, this drop can be collected directly from the pricked finger using the provided capillary. An additional 10µL of blood is collected using the second capillary (K_2_EDTA coated) and mixed with a diluent and dried fluorescent stains within the mixing-bottle. The resulting blood mixture is then used to fill the imaging chamber (Figure 1).

**Figure 1.**
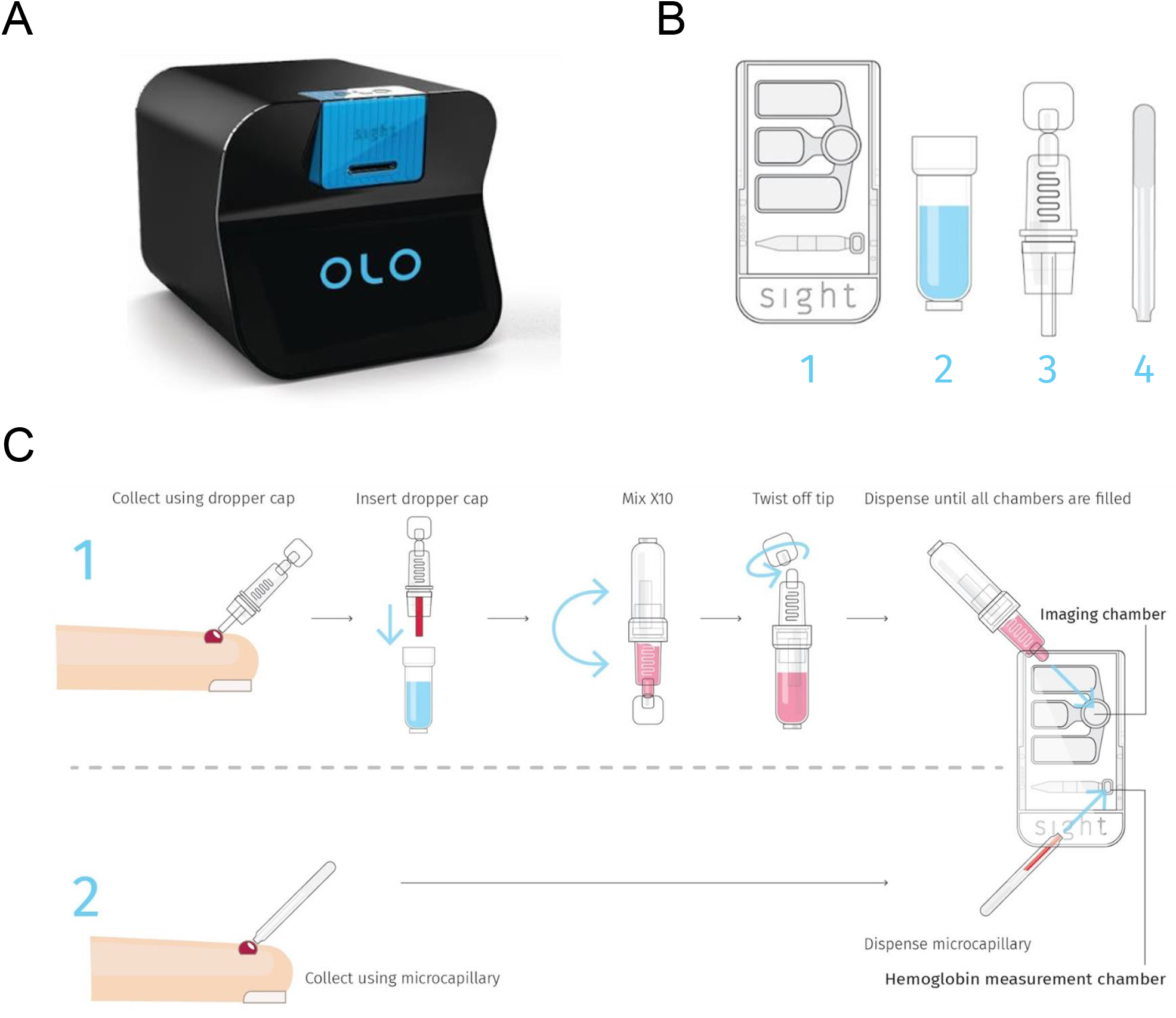
The Sight OLO hematology analyzer: (A) A rendering of the Sight OLO hematology analyzer (Sight Diagnostics, Israel), a desktop system measuring 284mm x 255mm x 324mm. (B) Components of OLO’s single-use test-kit: (1) cartridge, (2) mixing-bottle, (3) dropper-cap, (4) microcapillary. (C) Sample preparation workflow for finger-prick samples.

### Live Monolayer Imaging

To facilitate high-resolution microscopy of individual cells, the blood sample has to be presented for imaging as a monolayer - a film characterized by a defined focal plane in which cells rarely overlap with each other. Traditionally, this is accomplished through the preparation of blood smears; however, since the quality of blood films prepared using this process tends to vary significantly between users and across the sample area, the method was deemed unsuitable for POC usage.

Instead, Sight OLO relies on a novel monolayer formation process: the diluted blood sample is drawn using capillary action into the imaging chamber, which has a predefined height of a few hundred of microns. Since blood cells are denser than the diluent, they gradually sink and settle on the chamber floor. Due to the small height of the chamber, this process only takes about one minute, leaving the cells at a defined focal plane as desired for imaging. Furthermore, the dilution ratio is selected to ensure that, for the most part, the settled cells do not overlap regardless of the concentration of red blood cells in the sample (which ranges roughly between 2×10^6^/µL and 8×10^6^/µL). No sphering or fixation reagents are included in the diluent, in order to retain cell morphology.

### Staining and multispectral imaging

Traditionally, blood smears are stained with Wright-Giemsa stains and their variants, which have a century-long track-record in the differential identification of peripheral blood cell populations. However, these stains suffer from a number of drawbacks, including significant incubation times, the requirements for washing steps, the need for fixation (which disrupts morphology) and variability in stain appearance between preparations. These shortcomings prove challenging in the POC context, where speed, automation and a compact form-factor are desired. It is for this reason that Sight OLO departs from traditional staining methods and employs a novel staining and imaging approach.

Sight OLO uses a combination of brightfield and fluorescence microscopy, so that the identification of different cell populations can be aided by their optical and chemical signatures. In total, five illumination wavelengths are used to generate multispectral images of blood samples: violet (405nm), green (517nm) and red (633nm) brightfield channels as well as ultra-violet (365nm) and blue (460nm) fluorescent channels. Two fluorescent reagents are included in OLO’s sample preparation process and mixed with blood as it undergoes the dilution process. The first of these reagents stains cell nuclear DNA while the other stains DNA, RNA and acidic cell organelles. Additionally, violet illumination, which is highly absorbed by hemoglobin, assists in identifying RBCs and quantifying cellular hemoglobin content. Characteristic images of different normal and abnormal blood cell types as acquired by OLO are presented using false coloring in Figure 2.

**Figure 2.**
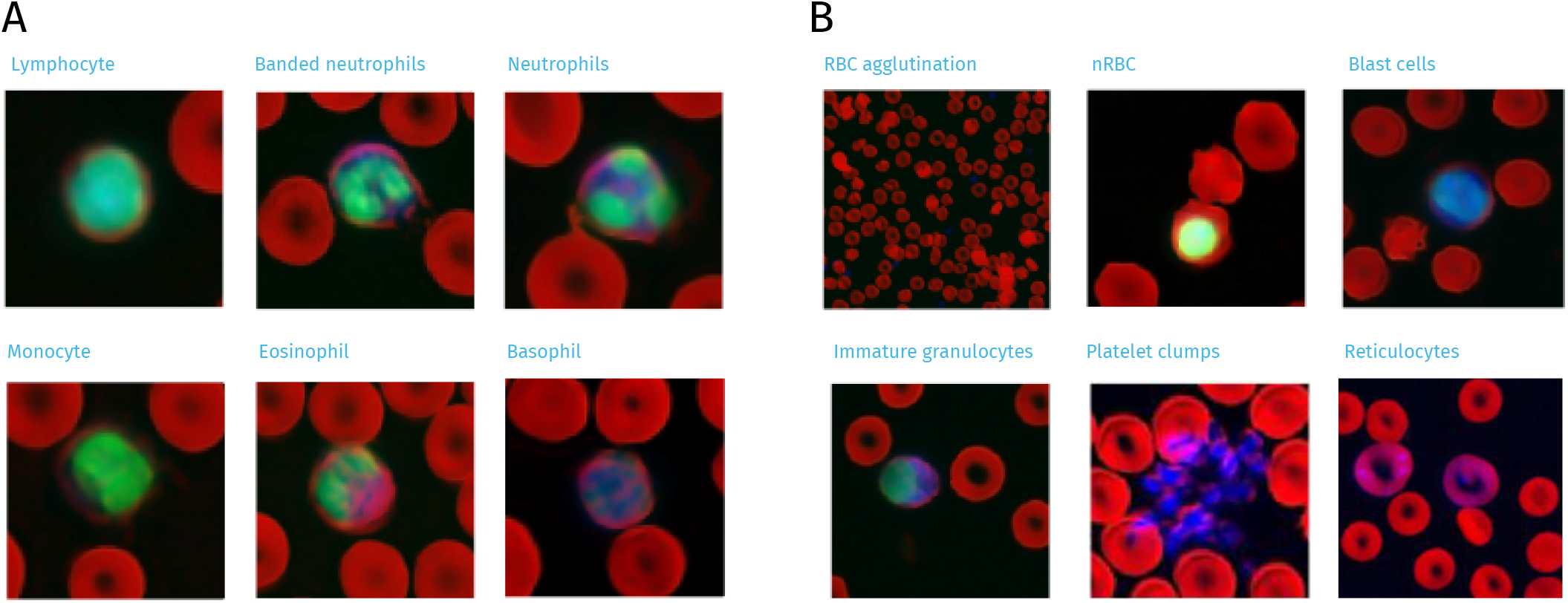
False-colored micrographs collected using OLO’s multispectral microscopy. Red channel: hemoglobin absorption; green channel: nuclear DNA fluorescence; blue channel: cytoplasmic staining. (A) characteristic examples of different white blood cell types. (B) characteristic examples of different anomalous cell types and formations.

### Scanning hardware

Sight OLO contains a fully automated fluorescence microscope which rapidly collects high-quality images of the blood smear present within its single-use test cartridge. The analyzer automatically scans at least 200 non-overlapping fields within the sample, and at each field it acquires the several channels of fluorescent and brightfield imaging. In order to simplify manufacturing constraints on cartridge flatness, OLO automatically refocuses on the sample at every field. Each blood sample is thus “digitized” into approximately 6 GB of image data.

In addition to microscopy, the Sight OLO also measures hemoglobin using an unlysed, reagent-free process utilizing four wavelength to account for absorption and scattering(13). The hemoglobin chamber is directly filled with undiluted whole blood using capillary action and contains several measurement areas, which differ by optical path length. These areas are used to derive differential measurements in order to normalize for system parameters such as illumination intensity and manufacturing tolerances.

### Algorithmic analysis

Sight OLO quantifies and characterizes cell populations in each image by employing three different analysis pipelines, one for each of the three primary cell types: red blood cells (RBCs), white blood cells (WBCs) and platelets. Each of these pipelines uses a similar two-stage process: first, “candidates” are generated based on certain characteristic features of the relevant cell population; then, each of these candidates is characterized using more in-depth analysis. Specifics for each of the three analysis pipelines are detailed below.

### Red Blood Cells

Red blood cells (RBC) are identified by the analyzer’s algorithms using brightfield images, which include images in the violet channel which is highly absorbed by the RBC hemoglobin content. This allows the detection algorithm to distinguish areas containing RBCs from their surrounding background. However, RBCs may still overlap due to landing too close to each other, an effect that can confound cell counts and per-cell measurements. Consequently, once candidates are identified, they are screened for overlaps and split into individual cells using morphological features as necessary for accurate counting using their morphological features.

RBCs are further characterized using Convolutional Neural network (CNN) algorithms in order to estimate cell properties such as Mean Cell Hemoglobin (MCH). Mean Cell Volume (MCV) and Mean Cell Hemoglobin Concentration (MCHC) are also obtained from the images.

### White Blood Cells

White blood cell (WBC) candidates are detected using the fluorescent staining exhibited by both their nucleus and cytoplasm, which must both be present. The candidates (which number in the order of a thousand in a typical sample) are then filtered according to size and shape to prevent debris and other unidentified objects from entering the classification process.

The two fluorescent channels together with the brightfield channels are used to classify each cell as a specific WBC type. To do so, each cell’s image is first segmented into a nucleus and a cytoplasm. It is then characterized by computing different features on either the cell, cytoplasm or nucleus; these features include morphological features (e.g. its area, diameter, etc.), intensity features (whether fluorescent emission or brightfield absorption) or texture (e.g. computing the standard deviation of the fluorescence of the nucleus and cytoplasm across their identified areas). These features are then used to classify the different WBC types, including abnormal cell types, by using machine-learning classification algorithms.

### Platelets

Platelets are detected using fluorescent staining. However, due to their low RNA content, platelets require a longer exposure time in order to obtain a strong enough signal to stand above the background. This signal is combined with the brightfield channels to detect the candidates. However, being much smaller and less bright than the other two cell types, cell fragments and debris are sometimes also detected as platelet candidates. Accordingly, true platelets are identified first by filtering the candidates according to different morphological and intensity properties, then applying several convolutional neural networks trained to accurately distinguish the platelets from background in different scenarios.

## RESULTS

We report here on the results of a clinical study conducted from June 2018 to March 2019 across three geographically diverse sites in the US: Boston Children’s Hospital (Boston, MA); Center for Advanced Laboratory Medicine, Columbia University (New York, NY); and TriCore Reference Laboratories (Albuquerque, NM). Some of the testing for the reproducibility and matrix comparison studies took place in Sight Diagnostics’ lab in Israel, with samples obtained from the Tel Aviv Sourasky Medical Center in Israel. The study protocol was reviewed and accepted by the Institutional Review Board (IRB) of each site separately.

The clinical and analytical performance assessments included method comparison with a comparative device, matrix comparison (venous vs. capillary and capillary vs. direct-from-finger samples), as well as repeatability, reproducibility and flagging studies.

## Method Comparison

The accuracy of Sight OLO was compared with the Sysmex XN-1000 System. The study design was based on the methods outlined below and in accordance with CLSI H20-A2(14), CLSI H26-A2(15)and CLSI EP09-A3(16). Samples from patients age 3 months to 94 years and included 355 males (52%) and 324 females (48%) were analyzed; 32% of the samples were from pediatric patients (3 months–21 years). Samples were selected to comprise several abnormalities, including different blood disorders and tumors (e.g. various types of anemias, Leukemias, Lymphomas, Myelomas), and covered a wide clinical range for each of the tested parameters.

The results of the regression analysis, which are included in Figure 3, show a strong concordance between the Sight OLO and the Sysmex XN both in terms of correlation coefficient and as seen through slope, bias and intercept. OLO performs with high accuracy for all CBC parameters. Detailed results are included in Supplemental Table 1. Supplemental Figure 1 and Supplemental Table 2 illustrate a version of the analysis without the exclusion of measurands invalidated by OLO, demonstrating the high number of actionable results.

**Figure 3.**
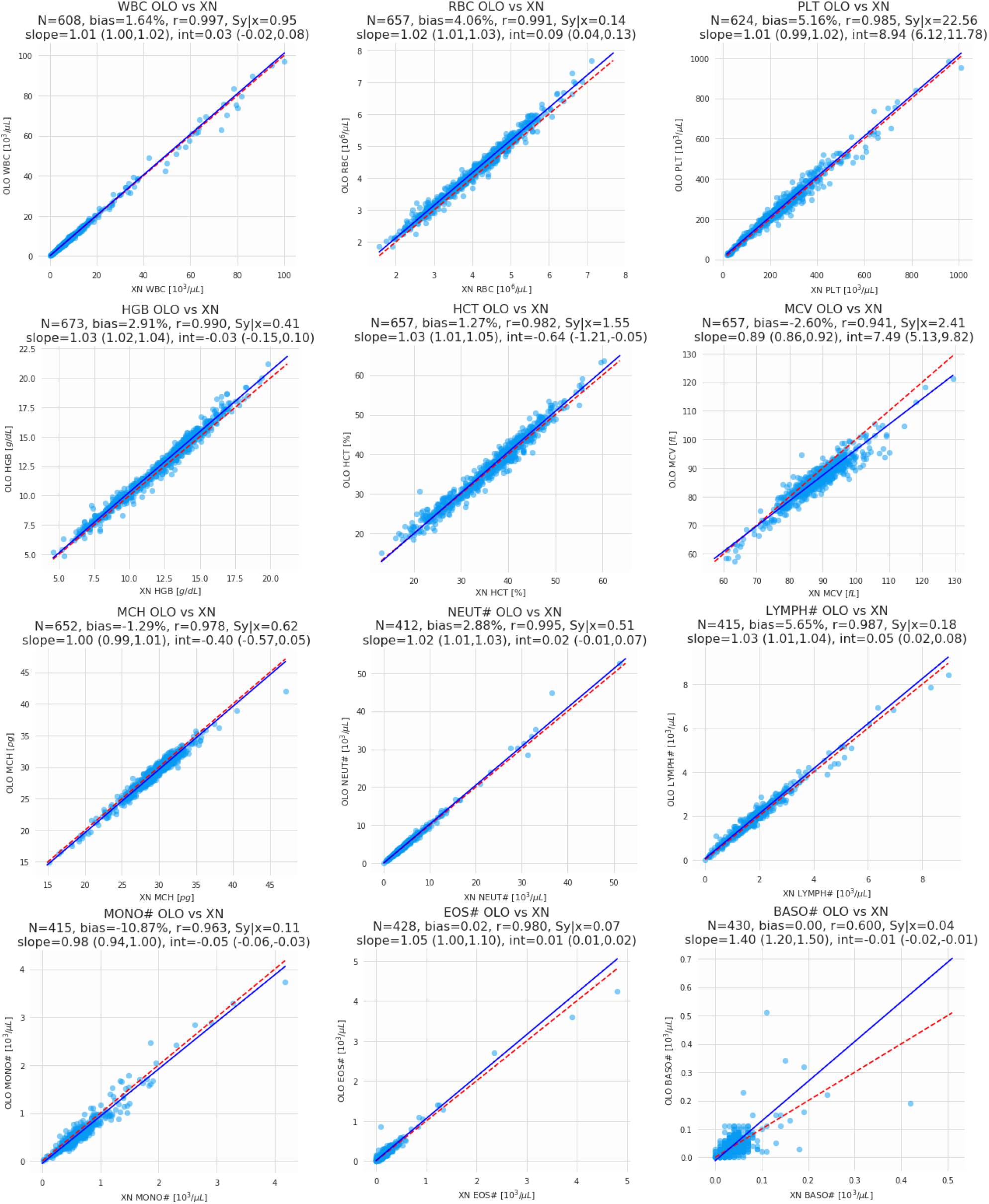
Results of the method comparison study between the Sight OLO and the Sysmex XN hematology analyzers. Graphs indicate Pearson correlation, slope and bias for each parameter. These results are tabulated in Supplemental Table 1.

### Repeatability and Reproducibility

Within-run repeatability studies were performed using residual K_2_EDTA whole blood samples as described below. In these studies, Sight OLO demonstrated high repeatability for most of the testing parameters. Table 1 shows the pooled SD for each measurand in relevant clinical ranges across all samples within that range. Reproducibility results are included in the Supplemental Table 4 and show similar performance to the within-run whole blood repeatability shown below.

**Table 1:** Whole Blood Repeatability Results.

| Pooled Results of Repeatability Study |  |  |  |  |  |  |
| --- | --- | --- | --- | --- | --- | --- |
| Measurand | Units | N Samples | N Replicates | Target Range | Mean | Pooled SD |
| WBC | 10<sup>3</sup>/μL | 4 | 78 | 0.5-4.0 | 2.04 | 0.12 |
| WBC | 10<sup>3</sup>/μL | 26 | 512 | 4.0-80.0 | 9.26 | 0.40 |
| RBC | 10<sup>6</sup>/μL | 4 | 80 | 1.0-3.5 | 3.02 | 0.06 |
| RBC | 10<sup>6</sup>/μL | 27 | 531 | 3.5-8.0 | 5.04 | 0.11 |
| PLT | 10<sup>3</sup>/μL | 7 | 136 | 20-150 | 78.52 | 4.76 |
| PLT | 10<sup>3</sup>/μL | 24 | 474 | 150-800 | 309.01 | 16.69 |
| HGB | g/dL | 7 | 139 | 5-11 | 8.95 | 0.2 |
| HGB | g/dL | 24 | 473 | 11-25 | 14.81 | 0.28 |
| HCT | % | 31 | 614 | 10-70 | 40.45 | 0.91 |
| MCV | fL | 28 | 554 | 50-150 | 85.28 | 0.58 |
| RDW | % | 28 | 552 | 10-40 | 15 | 0.34 |
| MCH | pg | 28 | 554 | 10-45 | 28.5 | 0.2 |
| MCHC | g/dL | 28 | 554 | 26-38 | 33.26 | 0.28 |
| NEUT% | % | 1 | 20 | WBC≤4.0x10<sup>3</sup>/μL | 56.97 | 1.78 |
| NEUT% | % | 20 | 398 | WBC>4.0x10<sup>3</sup>/μL | 66.82 | 1.41 |
| NEUT# | 10<sup>3</sup>/μL | 3 | 58 | WBC≤4.0x10<sup>3</sup>/μL | 0.97 | 0.09 |
| NEUT# | 10<sup>3</sup>/μL | 20 | 398 | WBC>4.0x10<sup>3</sup>/μL | 6.45 | 0.31 |
| LYMPH% | % | 1 | 20 | WBC≤4.0x10<sup>3</sup>/μL | 31.80 | 2.00 |
| LYMPH% | % | 20 | 398 | WBC>4.0x10<sup>3</sup>/μL | 22.62 | 1.26 |
| LYMPH# | 10<sup>3</sup>/μL | 3 | 58 | WBC≤4.0x10<sup>3</sup>/μL | 0.84 | 0.07 |
| LYMPH# | 10<sup>3</sup>/μL | 20 | 398 | WBC>4.0x10<sup>3</sup>/μL | 1.89 | 0.13 |
| MONO% | % | 1 | 58 | WBC≤4.0x10<sup>3</sup>/μL | 8.30 | 1.20 |
| MONO% | % | 20 | 398 | WBC>4.0x10<sup>3</sup>/μL | 7.74 | 0.86 |
| MONO# | 10<sup>3</sup>/μL | 3 | 58 | WBC≤4.0x10<sup>3</sup>/μL | 0.16 | 0.03 |
| MONO# | 10<sup>3</sup>/μL | 20 | 397 | WBC>4.0x10<sup>3</sup>/μL | 0.69 | 0.09 |
| EOS% | % | 1 | 58 | WBC≤4.0x10<sup>3</sup>/μL | 2.00 | 0.63 |
| EOS% | % | 20 | 398 | WBC>4.0x10<sup>3</sup>/μL | 2.29 | 0.49 |
| EOS# | 10<sup>3</sup>/μL | 3 | 58 | WBC≤4.0x10<sup>3</sup>/μL | 0.04 | 0.02 |
| EOS# | 10<sup>3</sup>/μL | 20 | 398 | WBC>4.0x10<sup>3</sup>/μL | 0.19 | 0.04 |
| BASO% | % | 1 | 58 | WBC≤4.0x10<sup>3</sup>/μL | 0.94 | 0.40 |
| BASO% | % | 20 | 398 | WBC>4.0x10<sup>3</sup>/μL | 0.54 | 0.25 |
| BASO# | 10<sup>3</sup>/μL | 3 | 58 | WBC≤4.0x10<sup>3</sup>/μL | 0.02 | 0.01 |
| BASO# | 10<sup>3</sup>/μL | 20 | 398 | WBC>4.0x10<sup>3</sup>/μL | 0.04 | 0.02 |

### White Blood Cell Flagging Study

The flagging capabilities of Sight OLO were compared to manual microscopy for WBC distributional abnormalities and WBC morphological abnormalities, which include blasts, immature granulocytes, nucleated RBCs, and atypical lymphocytes. Other invalidating messages, such as platelet clumps and RBC agglutination were not included in the flagging study. 108 negative and 100 positive samples with WBC count larger than 4×10^3^/µL were enrolled from the method comparison study. For each of the 208 samples, two qualified examiners evaluated the blood films and performed a 200-cell differential on one of three blood films, according to examination protocol. When abnormalities were present (e.g., distributional abnormalities based on manual differential count or morphological abnormality), the sample was considered abnormal. Otherwise, the sample was considered normal. Rumke analysis was used for ensuring agreements between the two examiners, whereby a third was used as an arbitrator if no such agreement was found. If no pair of examiners who agree with each other was found, the sample was discarded.

The flagging study tested for both WBC distributional abnormality and the following WBC morphology abnormalities that are flagged by Sight OLO: NRBCs, blast cells, immature granulocytes and atypical lymphocytes. OLO’s flagging was assessed for agreement with the assessment of the manual blood smears. The overall WBC flagging capabilities of OLO showed good clinical utility for both sensitivity and specificity, as seen in Supplemental Table 3. Specifically, the assessment of OLO’s flagging performance for the WBC abnormalities demonstrated sensitivity/positive percent agreement of 93% (95% CI: 86.1% - 97.1%), specificity/negative percent agreement of 80.6% (95% CI: 71.8% - 87.5%) and overall agreement of 86.5% (95% CI: 81.1% - 90.9%).

### Matrix Comparison

A matrix comparison study was performed in two parts to assess the equivalence of Sight OLO’s analysis between venous and capillary samples, and between capillary and direct-from-finger samples. The results of the comparison between the capillary and venous samples of 67 patients, including some outside of the normal range, is shown in Table 2. Similarly, a comparison between the 40 healthy patients’ direct-from-finger and capillary samples is shown in Table 3. In this analysis, a single sample was excluded as a clear monocyte outlier and was in fact invalidated for monocyte count and fraction. This sample was also abnormal, having less than 4×10^3^/µL Leukocytes. No other invalidated results were excluded from analysis. Both studies showed good agreement between the venous and capillary and between capillary and direct from finger-prick samples, with high correlation and small biases, supporting the use of the different specimen types for measurement on OLO.

**Table 2:**
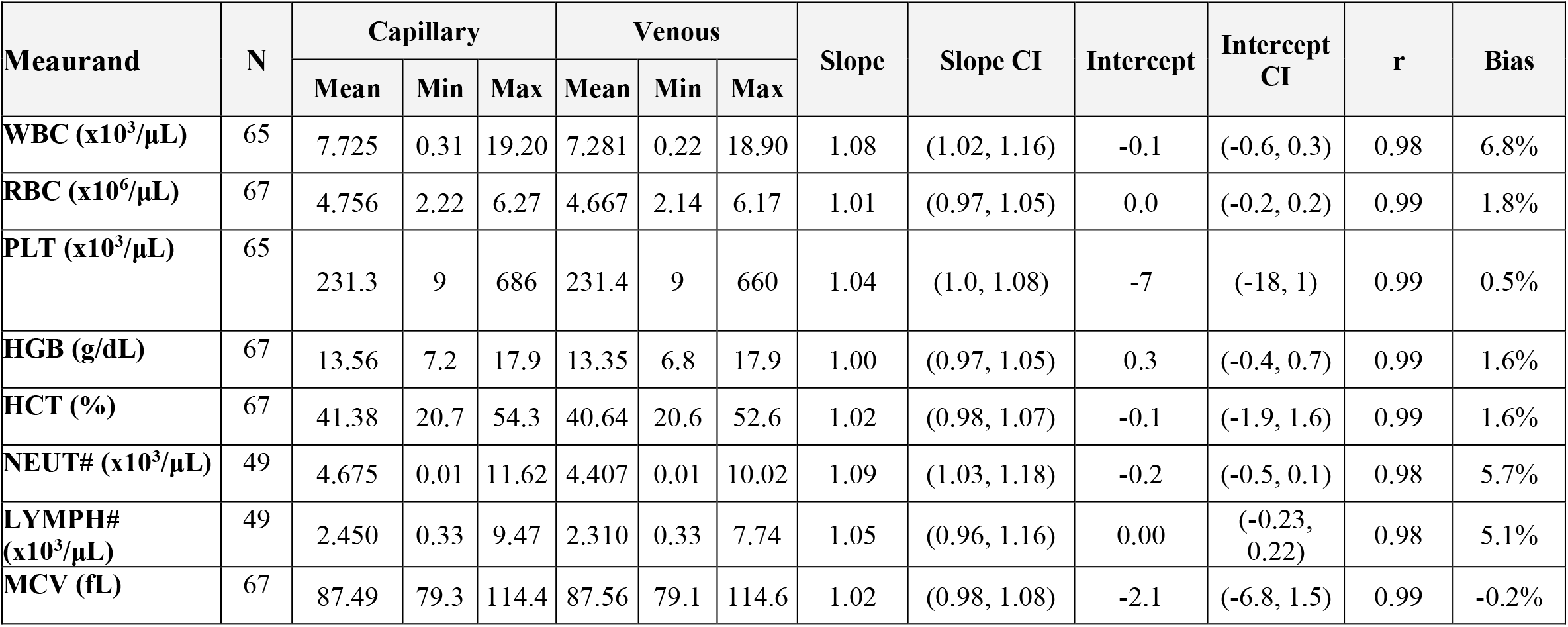
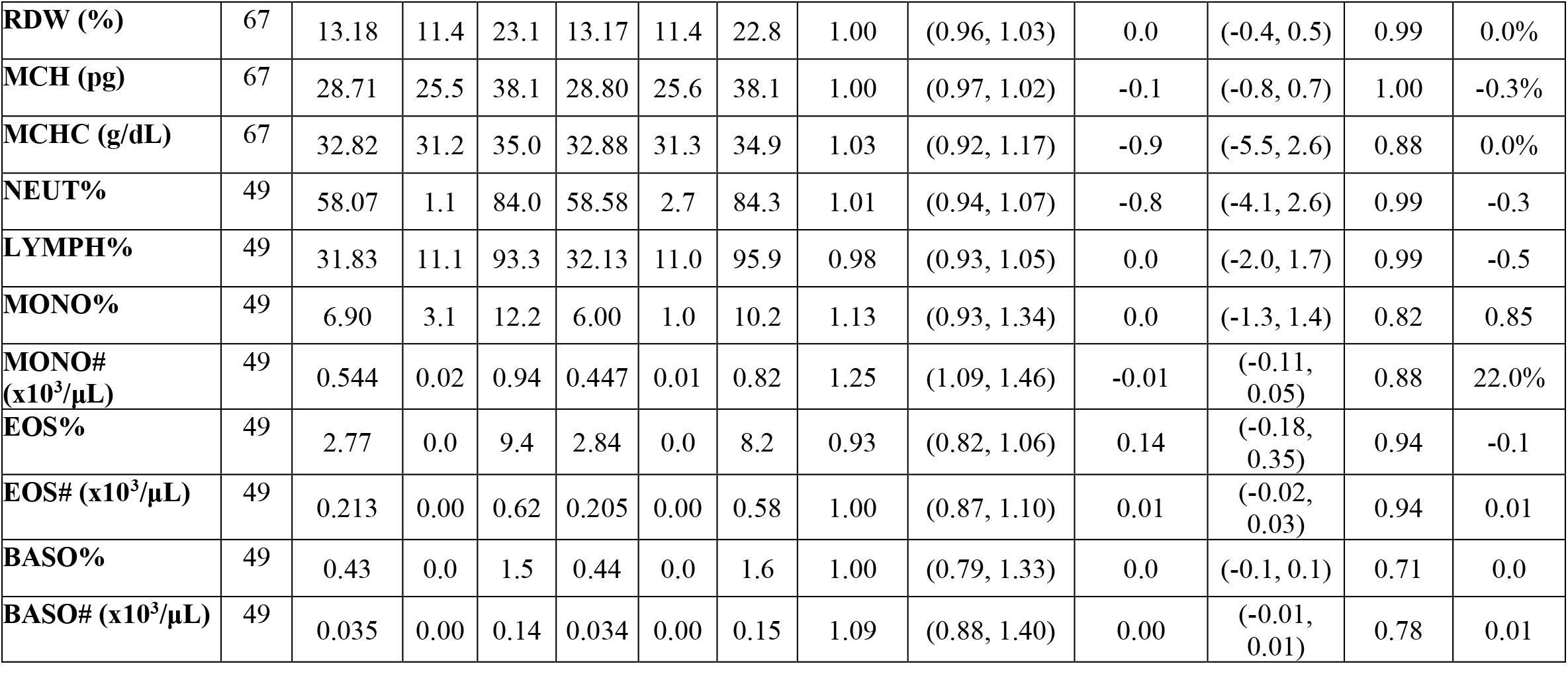
Venous vs. Capillary Matrix Study Results.

| Meaurand | N | Capillary |  |  | Venous |  |  | Slope | Slope CI | Intercept | Intercept CI | r | Bias |
| --- | --- | --- | --- | --- | --- | --- | --- | --- | --- | --- | --- | --- | --- |
|  |  | Mean | Min | Max | Mean | Min | Max |  |  |  |  |  |  |
| WBC ($\times 10^3/\mu\text{L}$) | 65 | 7.725 | 0.31 | 19.20 | 7.281 | 0.22 | 18.90 | 1.08 | (1.02, 1.16) | -0.1 | (-0.6, 0.3) | 0.98 | 6.8% |
| RBC ($\times 10^6/\mu\text{L}$) | 67 | 4.756 | 2.22 | 6.27 | 4.667 | 2.14 | 6.17 | 1.01 | (0.97, 1.05) | 0.0 | (-0.2, 0.2) | 0.99 | 1.8% |
| PLT ($\times 10^3/\mu\text{L}$) | 65 | 231.3 | 9 | 686 | 231.4 | 9 | 660 | 1.04 | (1.0, 1.08) | -7 | (-18, 1) | 0.99 | 0.5% |
| HGB (g/dL) | 67 | 13.56 | 7.2 | 17.9 | 13.35 | 6.8 | 17.9 | 1.00 | (0.97, 1.05) | 0.3 | (-0.4, 0.7) | 0.99 | 1.6% |
| HCT (%) | 67 | 41.38 | 20.7 | 54.3 | 40.64 | 20.6 | 52.6 | 1.02 | (0.98, 1.07) | -0.1 | (-1.9, 1.6) | 0.99 | 1.6% |
| NEUT# ($\times 10^3/\mu\text{L}$) | 49 | 4.675 | 0.01 | 11.62 | 4.407 | 0.01 | 10.02 | 1.09 | (1.03, 1.18) | -0.2 | (-0.5, 0.1) | 0.98 | 5.7% |
| LYMPH# ($\times 10^3/\mu\text{L}$) | 49 | 2.450 | 0.33 | 9.47 | 2.310 | 0.33 | 7.74 | 1.05 | (0.96, 1.16) | 0.00 | (-0.23, 0.22) | 0.98 | 5.1% |
| MCV (fL) | 67 | 87.49 | 79.3 | 114.4 | 87.56 | 79.1 | 114.6 | 1.02 | (0.98, 1.08) | -2.1 | (-6.8, 1.5) | 0.99 | -0.2% |
| **RDW (%)** | 67 | 13.18 | 11.4 | 23.1 | 13.17 | 11.4 | 22.8 | 1.00 | (0.96, 1.03) | 0.0 | (-0.4, 0.5) | 0.99 | 0.0% |
| **MCH (pg)** | 67 | 28.71 | 25.5 | 38.1 | 28.80 | 25.6 | 38.1 | 1.00 | (0.97, 1.02) | -0.1 | (-0.8, 0.7) | 1.00 | -0.3% |
| **MCHC (g/dL)** | 67 | 32.82 | 31.2 | 35.0 | 32.88 | 31.3 | 34.9 | 1.03 | (0.92, 1.17) | -0.9 | (-5.5, 2.6) | 0.88 | 0.0% |
| **NEUT%** | 49 | 58.07 | 1.1 | 84.0 | 58.58 | 2.7 | 84.3 | 1.01 | (0.94, 1.07) | -0.8 | (-4.1, 2.6) | 0.99 | -0.3 |
| **LYMPH%** | 49 | 31.83 | 11.1 | 93.3 | 32.13 | 11.0 | 95.9 | 0.98 | (0.93, 1.05) | 0.0 | (-2.0, 1.7) | 0.99 | -0.5 |
| **MONO%** | 49 | 6.90 | 3.1 | 12.2 | 6.00 | 1.0 | 10.2 | 1.13 | (0.93, 1.34) | 0.0 | (-1.3, 1.4) | 0.82 | 0.85 |
| **MONO#**<br>**(x10<sup>3</sup>/μL)** | 49 | 0.544 | 0.02 | 0.94 | 0.447 | 0.01 | 0.82 | 1.25 | (1.09, 1.46) | -0.01 | (-0.11, 0.05) | 0.88 | 22.0% |
| **EOS%** | 49 | 2.77 | 0.0 | 9.4 | 2.84 | 0.0 | 8.2 | 0.93 | (0.82, 1.06) | 0.14 | (-0.18, 0.35) | 0.94 | -0.1 |
| **EOS# (x10<sup>3</sup>/μL)** | 49 | 0.213 | 0.00 | 0.62 | 0.205 | 0.00 | 0.58 | 1.00 | (0.87, 1.10) | 0.01 | (-0.02, 0.03) | 0.94 | 0.01 |
| **BASO%** | 49 | 0.43 | 0.0 | 1.5 | 0.44 | 0.0 | 1.6 | 1.00 | (0.79, 1.33) | 0.0 | (-0.1, 0.1) | 0.71 | 0.0 |
| **BASO# (x10<sup>3</sup>/μL)** | 49 | 0.035 | 0.00 | 0.14 | 0.034 | 0.00 | 0.15 | 1.09 | (0.88, 1.40) | 0.00 | (-0.01, 0.01) | 0.78 | 0.01 |

**Table 3:** Finger-Prick vs. Capillary Matrix Study Results

| Measurand | N | Finger |  |  | Capillary |  |  | Slope | Slope CI | Intercept | Intercept CI | r | Bias |
| --- | --- | --- | --- | --- | --- | --- | --- | --- | --- | --- | --- | --- | --- |
|  |  | Mean | Min | Max | Mean | Min | Max |  |  |  |  |  |  |
| **WBC**<br>(x10<sup>3</sup>/μL) | 40 | 8.08 | 3.75 | 12.16 | 8.01 | 3.8 | 12.51 | 0.97 | (0.89, 1.06) | 0.3 | (-0.3, 0.9) | 0.97 | 1.4% |
| **RBC**<br>(x10<sup>6</sup>/μL) | 40 | 4.797 | 3.61 | 6.14 | 5.081 | 4.07 | 6.23 | 0.99 | (0.87, 1.10) | -0.2 | (-0.8, 0.4) | 0.96 | -5.3% |
| **PLT**<br>(x10<sup>3</sup>/μL) | 40 | 262.9 | 138 | 431 | 264.2 | 148 | 412 | 1.02 | (0.87, 1.17) | -7 | (-44, 36) | 0.92 | -0.7% |
| **HGB**<br>(g/dL) | 40 | 13.56 | 10.5 | 17.5 | 14.35 | 12.0 | 17.8 | 1.02 | (0.90, 1.16) | -1.0 | (-3.1, 0.7) | 0.95 | -5.2% |
| **MCV**<br>(fL) | 40 | 84.43 | 77.5 | 93.4 | 84.35 | 78.2 | 92.6 | 1.01 | (0.89, 1.10) | -0.8 | (-7.8, 9.6) | 0.95 | 0.3% |
| **RDW**<br>(%) | 40 | 12.58 | 11.2 | 13.7 | 12.48 | 11.2 | 14.1 | 0.94 | (0.73, 1.13) | 0.9 | (-1.5, 3.4) | 0.85 | 1.2% |
| **MCH**<br>(pg) | 40 | 28.31 | 25.2 | 31.6 | 28.28 | 25.2 | 31.4 | 1.00 | (0.95, 1.02) | 0.1 | (-0.4, 1.4) | 1.00 | 0.2% |
| **MCHC**<br>(g/dL) | 40 | 33.54 | 31.7 | 36.2 | 33.54 | 31.1 | 35.8 | 1.01 | (0.89, 1.17) | -0.5 | (-5.7, 3.8) | 0.92 | 0.0% |
| **NEUT#**<br>(x10<sup>3</sup>/μL) | 40 | 4.53 | 1.61 | 7.66 | 4.58 | 1.75 | 8.56 | 0.96 | (0.9, 1.06) | 0.1 | (-0.3, 0.4) | 0.97 | -1.4% |
| **LYMPH#**<br>(x10<sup>3</sup>/μL) | 40 | 2.74 | 1.58 | 5.21 | 2.58 | 1.4 | 3.94 | 1.04 | (0.93, 1.16) | 0.07 | (-0.23, 0.34) | 0.95 | 7.3% |
| **NEUT%** | 40 | 55.16 | 39.8 | 70.5 | 56.35 | 38.1 | 72 | 1.06 | (0.96, 1.17) | -4.5 | (-10.9, 1.5) | 0.95 | -1.1 |
| **LYMPH%** | 40 | 34.64 | 21.5 | 49.4 | 32.89 | 20.2 | 50.2 | 1.03 | (0.94, 1.13) | 0.7 | (-2.8, 3.4) | 0.96 | 1.6 |
| **MONO%** | 39 | 7.16 | 3.0 | 12.7 | 7.81 | 4.6 | 13.6 | 0.88 | (0.70, 1.11) | 0.2 | (-1.4, 1.6) | 0.80 | -0.6 |
| **MONO#**<br>(x10<sup>3</sup>/μL) | 39 | 0.57 | 0.22 | 1.13 | 0.62 | 0.31 | 0.93 | 0.93 | (0.73, 1.20) | 0.00 | (-0.16, 0.12) | 0.74 | -7.8% |
| **EOS%** | 40 | 2.56 | 0.1 | 13 | 2.54 | 0.2 | 12.1 | 1.01 | (0.92, 1.13) | 0.0 | (-0.2, 0.2) | 0.97 | 0.1 |
| **EOS#**<br>(x10<sup>3</sup>/μL) | 40 | 0.21 | 0.01 | 1.3 | 0.2 | 0.02 | 1.09 | 1.04 | (0.92, 1.13) | 0.00 | (-0.01, 0.02) | 0.98 | 0.01 |
| **BASO%** | 40 | 0.39 | 0 | 1.3 | 0.46 | 0 | 1.4 | 0.75 | (0.57, 1.0) | 0.0 | (-0.0, 0.1) | 0.74 | 0.0 |
| **BASO#**<br>(x10<sup>3</sup>/μL) | 40 | 0.03 | 0 | 0.09 | 0.04 | 0 | 0.11 | 0.83 | (0.67, 1.0) | 0.00 | (-0.01, 0.00) | 0.68 | -0.01 |

## DISCUSSION

This multicenter evaluation provides a comprehensive validation of the performance characteristics of the Sight OLO hematological platform. Method comparison, repeatability and reproducibility studies demonstrate that the OLO analyzer provides CBC results that are comparable with the Sysmex XN across a wide measuring range, including highly challenging samples. A high correlation (r*≥*0.94) was found between most RBC parameters and WBC differentials (Figure 3). In the differential, a very high correlation (r>0.98) was seen for neutrophils, lymphocytes and eosinophils whereas a moderately high correlation (r=0.89) was found for monocytes fraction and a r=0.67 correlation was found for basophils faction. Also, low correlation was found for mean corpuscular hemoglobin concentration (r=0.69). The slightly low correlation for basophils is not uncommon, as the typical data range for this parameter is very limited, leading to low correlation values. The slightly low correlation for MCHC is due to the fact that in both OLO and the Sysmex XN, MCHC is a calculated parameter based on the ratio of two highly correlated values (MCH and MCV or HGB and HCT), leading to a small range of MCHC values; such ratios mathematically amplify any small system-specific differences in either of the parameters, thereby harming the correlation in the calculated parameter despite the very high correlation of MCH and MCV. These findings are equivalent to those observed for comparison between other modern laboratory analyzers(11, 17, 18). The OLO reproducibility and repeatability values were found to be within the CLIA allowable total errors for the relevant measurands. The present study also evaluated OLO’s automated flagging of samples for the presence of distributional abnormalities, NRBCs, blast cells, immature granulocytes andatypical lymphocytes. Results indicate that for these flags the OLO system was in high accordance with manual microscopy.

Sight OLO offers a full CBC utilizing only 27µL of blood sample, providing substantial blood conservation for oncology, neonatal and pediatric patients(19). While OLO is currently indicated for use in patients 3 months and older, further studies are currently being conducted to increase the population range. Furthermore, the fact that 27µL may be directly drawn from a finger-prick reduces some of the additional overheads and inconveniences associated with venous phlebotomy or larger volume capillary collection and may alleviate the anxiety involved with these for certain apprehensive populations such as pediatrics or patients who poorly tolerate traditional needle-based blood drawings. The disposable cartridge utilized by OLO has added benefits, as the risk of carryover or system clogging are eliminated, and no maintenance and cleaning are required between runs.

The study presented here successfully validated the performance of Sight OLO hematology analyzer in a multicenter clinical laboratory setting. In particular, the study demonstrated that OLO is accurate and comparable to the renowned Sysmex XN Series Hematology Analyzers. This study led to OLO’s FDA 510(k) clearance(12), and it demonstrates the capabilities of multi-spectral live monolayer imaging and AI-assisted image analysis in hematology.

The study also demonstrated the validity of performing 5-part differential CBC analysis using direct-from-finger blood samples: Sight OLO was able to produce results from fingerpricks that are equivalent to those obtained using venous blood draws. Of note, OLO is the first CBC analyzer to be cleared by the FDA for collecting samples directly from fingerpicks.

## MATERIALS AND METHODS

### Clinical protocol

#### Method comparison study

A method comparison study was undertaken to compare Sight OLO with the Sysmex XN-1000 System. The study design was based on the methods outlined in CLSI H20-A2(14), CLSI H26-A2(15) and CLSI EP09-A3(16), and is consistent with the approach conducted for FDA submissions for hematology devices.

Residual whole blood clinical samples (N = 679) were collected from both adult (>22 years old, N = 462) and pediatric patients (3 months to 21 years old, N= 217). All samples were collected in standard K_2_EDTA collection tubes (Becton Dickinson, Franklin Lakes, NJ, USA) and processed within 8 hours of venipuncture. Processing included testing on both analyzers within 2 hours of each other, as well as preparing three blood smears for each sample. The study included both normal and pathological samples in order to assess Sight OLO’s performance across the analytical measuring range and around medical decision points.

A Passing-Bablok regression analysis was performed for each CBC parameter after excluding any result which is invalidated by Sight OLO or by the comparative method. For each regression analysis, the slope and intercept and the 95% two-sided confidence interval (CI) around the slope and intercept were calculated, as well as the correlation coefficient. The overall bias was calculated as the median of differences, where differences were taken either as absolute or relative, according to the nature of the data.

#### Repeatability study

Within-run repeatability studies were performed using residual K_2_EDTA whole blood samples (minimum of 2mL per sample). In order to span healthy and pathological values and the medical decision points between them, each site tested at least 4 samples within lab reference ranges, 3 samples around medical decision levels for HGB (6-10g/dL), PLT(<50×10^3^/µL) and WBC(<2×10^3^/µL), and 4 samples around the upper range for RBC(>6×10^6^/µL), HGB(>17 g/dL), WBC(>12×10^3^/µL) and PLT(>600×10^3^/µL). In total, this requirement led to 38 samples being scanned, with each sample measured 20 consecutive times (after excluding invalidations or rejects). Standard deviation (SD) and coefficient of variation were calculated for each run. The first 20 successful runs per measurand were analyzed. If fewer than 20 successful scans were obtained within the required time slot from phlebotomy (8h), the sample was still analyzed so long as 17 or more replicates were scanned. For the anemic samples (HGB 6-10 g/dL) only RBC, HGB and HCT were analyzed, while for the thrombocytopenic(<50×10^3^/µL) and leukopenic (<2×10^3^/µL) samples, only PLT and only the WBC concentration and differential were analyzed, respectively. This was in accordance with the CLSI guidance H26-A2(15), which refers to the ICSH protocol for evaluation of blood cell counters.

#### Reproducibility studies

Reproducibility studies were conducted using three levels of commercial control materials (low, normal and high - below, within and above the reference ranges of main parameters, respectively) for all reported parameters. A total of 240 samples were included for each level of control. Controls (R&D Systems, Minneapolis, MN) were measured over 5 operating days at three sites (Boston Children’s Hospital, Columbia University and Sight’s laboratory), with two devices at each site for a total of six devices. Two runs per day, and four replicates per run were performed. At each site the testing was performed by two operators, where the first operator conducted the first run of all days and the second operator conducted the second run. Every replicate required a new test kit and, every day, a new quality control material tube per level was opened at each site. This was in accordance with the CLSI guideline EP-05-A3.

#### Flagging study

The flagging capabilities of Sight OLO were compared to manual microscopy for WBC distributional abnormalities and WBC morphological abnormalities, which include blasts, immature granulocytes, nucleated RBCs, and atypical lymphocytes. Other invalidating messages, such as platelet clumps and RBC agglutination were not included in the flagging study. 108 negative and 100 positive samples with WBC count larger than 4×10^3^/µL were enrolled from the method comparison study. For each sample, the three prepared smears were sent to analysis by trained morphologists from Columbia University Irving Medical Center staff, who had no access to either clinical information or reference method results. The testing design was based on the test methods outlined in CLSI H20-A2(14) and CLSI H26-A2(15).

#### Matrix comparison study

A matrix comparison study was performed in two parts to assess the equivalence between venous and capillary samples, and between capillary and direct-from-finger samples. In the first part of the study, 67 samples (of 52 subjects, 15 of which repeated the test after a six-month interval) were collected in pairs. A venous sample was drawn, and a capillary sample was collected into 350uL microtainers. Healthy volunteers were primarily tested, and additional samples at medical decision points and across the analytical measuring range (i.e.: HGB: 6-10 g/dL; WBC<2×10^3^/µL; PLT<50×10^3^/µL; WBC>12×10^3^/µL; PLTs>500×10^3^/µL; RBCs>6×10^6^/µL; HGB>17g/dL) were enrolled from subjects in Tel Aviv Sourasky Medical Center. Each pair of samples was tested on the same Sight OLO device 4 times: two replicates of the venous sample and two of the capillary samples. After excluding invalidated results, a Passing-Bablok regression analysis was performed between the pair-averaged capillary and pair-averaged venous samples.

For the second part of the matrix-comparison study, 40 apparently healthy patients were enrolled, and capillary samples were collected from each of them using two different methods. First, a capillary sample was collected into a 350uL microtainer. Then, two finger-prick samples from different fingers were collected, 27uL from each, directly into the Sight OLO test kit’s microcapillaries and immediately processed on the OLO. The two finger-prick samples and the two repeats of the capillary sample collected into the microtainer were averaged, and results were compared to each other using Passing-Bablok analysis.

## Data Availability

Data and analysis code is available on request from the authors and will be shared under a MTA.

## Research Funding

S.D. Sight Diagnostics LTD

## Author Contributions

All authors confirm their contribution to the intellectual content of this manuscript. All authors have met the following requirements: a) significant contribution to conception and design, data acquisition and/or analysis, and data interpretation; b) drafting or revision of the intellectual content of the article; c) final approval of the article if published.

Conceptualization: YE, DL, AZ, SL

Methodology: DB, SP, DB, EY, NB, SL

Investigation: DB, SP, DB, EY, NB, SL CS, AK, CB

Supervision: SL, CS, AK, CB

Writing – original draft: YA, DL, AZ, CB

Writing – review & editing: YA, DL, AZ, CS, AK, CB

## Competing Interests

*Employment or Leadership*: Y. Eshel, S.D. Sight Diagnostics LTD; D. Levner, S.D. Sight Diagnostics LTD; A. Zait, S.D. Sight Diagnostics LTD; D. Glück, S.D. Sight Diagnostics LTD; D. Benbassat, S.D. Sight Diagnostics LTD; S. Pecker, S.D. Sight Diagnostics LTD; D. Brailovsky, S.D. Sight Diagnostics LTD; E. Yurkovsky, S.D. Sight Diagnostics LTD; N. Bachar, S.D. Sight Diagnostics LTD; S. Levy, S.D. Sight Diagnostics LTD.

*Consultant or Advisory Role*: None declared

*Stock Ownership*: Y. Eshel, D. Levner, A. Zait, D. Glück, D. Benbassat, S. Pecker, D. Brailovsky, E. Yurkovsky, N. Bachar, and S. Levy have ownership of S.D. Sight Diagnostics LTD stocks or stock options.

*Honoraria*: None Declared

*Expert Testimony*: None declared

*Patents*: Y. Eshel, D. Levner, A. Zait, D. Glück, D. Benbassat, S. Pecker, D. Brailovsky, and S. Levy are inventors of patent applications and issued patents assigned to S.D. Sight Diagnostics LTD (PCT/IL2017/050526; PCT/IL2017/050523; PCT/IB2020/061728; PCT/IB2020/061724; PCT/IL2016/051025; PCT/IB2020/061732; PCT/IB2020/061736 PCT/IB2020/061731; PCT/IB2020/059924).

*Other remunerations*: None declared

*Role of Sponsor*: The funding organization played a direct role in study design, data review and interpretation, manuscript preparation, and manuscript final approval.

## Data and materials availability

data and analysis code is available on request from the authors and will be shared under a MTA.

## Supplementary materials and methods

**Supplemental Table S1:** Summary of the method comparison study comparing the Sight OLO and the Sysmex XN and presented in Figure 3.

| Measurand | N | Results Range | Correlation Coefficient (r) | Slope (95% CI) | Intercept (95% CI) | Median Bias | Median Relative Bias (%) |
| --- | --- | --- | --- | --- | --- | --- | --- |
| WBC $\times 10^3/\mu\text{L}$ | 608 | 0.30 to 97.16 | 0.997 | 1.011 (1.003, 1.020) | 0.033 (-0.016, 0.082) | 0.1 | 1.6 |
| RBC $\times 10^6/\mu\text{L}$ | 657 | 1.86 to 7.69 | 0.991 | 1.019 (1.008, 1.030) | 0.089 (0.043, 0.129) | 0.16 | 4.1 |
| PLT $\times 10^3/\mu\text{L}$ | 624 | 22 to 985 | 0.985 | 1.006 (0.994, 1.021) | 8.937 (6.103, 11.801) | 10 | 5.2 |
| HGB g/dL | 673 | 4.9 to 21.2 | 0.990 | 1.031 (1.020, 1.042) | -0.028 (-0.150, 0.104) | 0.3 | 2.9 |
| HCT | 657 | 15.2 to 63.7 | 0.982 | 1.030 (1.014, 1.045) | -0.639 (-1.214, -0.044) | 0.4 | 1.3 |
| MCV fL | 657 | 57.3 to 121.2 | 0.941 | 0.889 (0.862, 0.916) | 7.489 (5.119, 9.821) | -2.3 | -2.6 |
| RDW | 647 | 10.6 to 29.4 | 0.939 | 1.000 (0.980, 1.036) | -0.100 (-0.607, 0.171) | -0.1 | -0.7 |
| MCH pg | 652 | 14.9 to 42.0 | 0.978 | 1.000 (0.986, 1.007) | -0.400 (-0.583, 0.046) | -0.4 | -1.3 |
| MCHC g/dL | 652 | 26.0 to 36.6 | 0.693 | 0.667 (0.625, 0.731) | 11.567 (9.431, 12.944) | 0.5 | 1.5 |
| NEUT% | 419 | 0.4 to 96.4 | 0.989 | 0.994 (0.980, 1.008) | 0.980 (0.051, 1.861) | 0.5 | 0.9 |
| NEUT# $\times 10^3/\mu\text{L}$ | 412 | 0.01 to 52.59 | 0.995 | 1.023 (1.011, 1.033) | 0.020 (-0.014, 0.070) | 0.12 | 2.9 |
| LYMPH% | 423 | 0.9 to 99.6 | 0.992 | 1.000 (0.989, 1.013) | 0.800 (0.452, 1.033) | 0.8 | 3.3 |
| LYMPH# $\times 10^3/\mu\text{L}$ | 415 | 0.02 to 8.44 | 0.987 | 1.025 (1.007, 1.044) | 0.050 (0.022, 0.083) | 0.09 | 5.7 |
| MONO% | 423 | 0.0 to 23.4 | 0.886 | 0.962 (0.910, 1.000) | -0.646 (-0.900, -0.264) | -0.9 | -12.5 |
| MONO# $\times 10^3/\mu\text{L}$ | 415 | 0.00 to 3.74 | 0.964 | 0.982 (0.941, 1.000) | -0.049 (-0.060, -0.027) | -0.06 | -10.9 |
| EOS% | 436 | 0.0 to 33.3 | 0.979 | 1.023 (1.000, 1.071) | 0.170 (0.100, 0.200) | 0.2 | 11.1 |
| EOS# $\times 10^3/\mu\text{L}$ | 427 | 0.00 to 4.24 | 0.986 | 1.042 (1.000, 1.094) | 0.014 (0.009, 0.020) | 0.02 | 15.0 |
| BASO% | 438 | 0.0 to 3.1 | 0.670 | 1.500<br>(1.333, 1.667) | -0.250<br>(-0.300, -0.167) | 0 | 0.0 |
| BASO#<br>x10<sup>3</sup>/μL | 429 | 0.00 to 0.34 | 0.641 | 1.333<br>(1.200, 1.500) | -0.010<br>(-0.015, -0.006) | 0 | 0.0 |

BASO, basophil; CI, confidence interval; EOS, eosinophil; HCT, hematocrit; HGB, hemoglobin; LYMPH, lymphocyte; MCHC, mean corpuscular hemoglobin concentration; MCH, mean corpuscular hemoglobin; MCV, mean corpuscular volume; MONO, monocyte; NEUT, neutrophil; PLT, platelet; WBC, white blood cell; RBC, red blood cell; RDW, red blood cell distribution width.

**Supplemental Figure S1.**
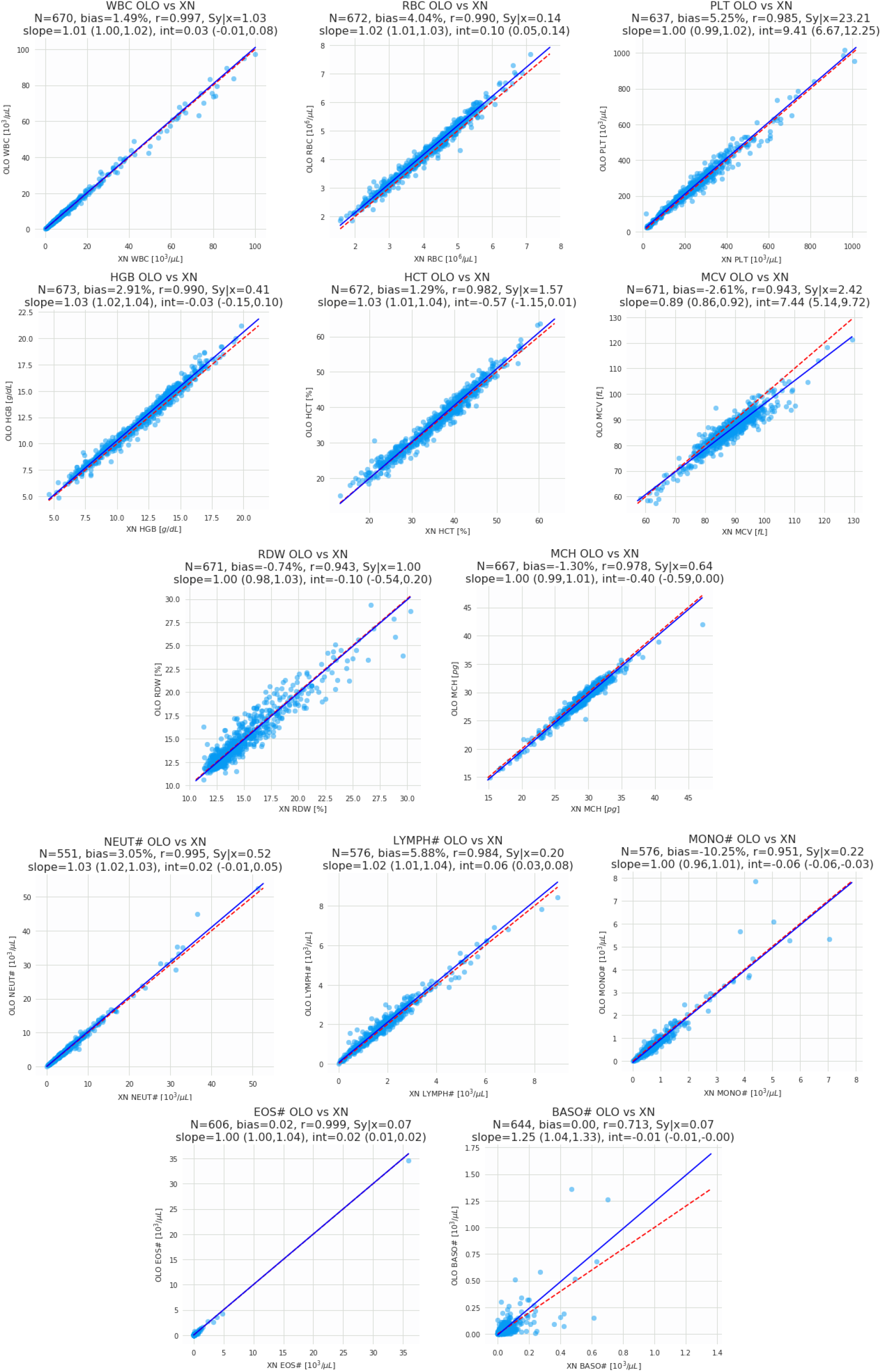
Results of the method comparison study between the Sight OLO and the Sysmex XN hematology analyzers with measurands invalidated by the OLO included in the analysis (while measurands invalidated by the Sysmex still excluded). Graphs indicate Pearson correlation, slope and bias for each parameter. These results are tabulated in Supplemental Table 3.

**Supplemental Table S2:** Summary of the method comparison study including results invalidated by OLO (corresponding to Supplemental Figure 1).

| Measurand | N | Results Range | Correlation Coefficient (r) | Slope (95% CI) | Intercept (95% CI) | Median Bias | Median Relative Bias (%) |
| --- | --- | --- | --- | --- | --- | --- | --- |
| WBC x10<sup>3</sup>/μL | 670 | 0.3 to 97.16 | 0.997 | 1.009 (1.001, 1.018) | 0.034 (-0.012, 0.083) | 0.09 | 1.5 |
| RBC x10<sup>6</sup>/μL | 672 | 1.86 to 7.69 | 0.99 | 1.017 (1.006, 1.027) | 0.098 (0.053, 0.139) | 0.16 | 4.0 |
| PLT x10<sup>3</sup>/μL | 637 | 22.0 to 1017.0 | 0.985 | 1.004 (0.992, 1.018) | 9.413 (6.67, 12.248) | 10 | 5.2 |
| HGB g/dL | 673 | 4.9 to 21.2 | 0.99 | 1.031 (1.02, 1.042) | -0.028 (-0.15, 0.104) | 0.3 | 2.9 |
| HCT | 672 | 15.2 to 63.7 | 0.982 | 1.028 (1.012, 1.044) | -0.567 (-1.152, 0.015) | 0.4 | 1.3 |
| MCV fL | 671 | 57.3 to 121.2 | 0.943 | 0.889 (0.863, 0.915) | 7.444 (5.142, 9.725) | -2.3 | -2.6 |
| RDW | 671 | 10.6 to 29.4 | 0.943 | 1.0 (0.979, 1.031) | -0.1 (-0.537, 0.196) | -0.1 | -0.7 |
| MCH pg | 667 | 14.9 to 42.0 | 0.978 | 1.0 (0.987, 1.007) | -0.4 (-0.592, 0.004) | -0.4 | -1.3 |
| MCHC g/dL | 667 | 26.0 to 37.2 | 0.7 | 0.683 (0.633, 0.738) | 10.998 (9.193, 12.66) | 0.5 | 1.5 |
| NEUT% | 551 | 0.4 to 96.4 | 0.987 | 0.995 (0.982, 1.008) | 1.029 (0.136, 1.841) | 0.7 | 1.0 |
| NEUT# x10<sup>3</sup>/μL | 551 | 0.01 to 52.59 | 0.995 | 1.025 (1.016, 1.034) | 0.016 (-0.01, 0.052) | 0.12 | 3.0 |
| LYMPH% | 576 | 0.9 to 99.6 | 0.99 | 1.0 (0.988, 1.009) | 0.8 (0.588, 1.083) | 0.8 | 3.8 |
| LYMPH# x10<sup>3</sup>/μL | 576 | 0.02 to 8.44 | 0.984 | 1.022 (1.006, 1.038) | 0.056 (0.034, 0.084) | 0.09 | 5.9 |
| MONO% | 576 | 0.0 to 37.1 | 0.925 | 0.917 (0.879, 0.957) | -0.221 (-0.526, 0.088) | -0.9 | -10.9 |
| MONO# x10<sup>3</sup>/μL | 576 | 0.0 to 7.86 | 0.951 | 1.0 (0.958, 1.008) | -0.06 (-0.062, -0.032) | -0.06 | -10.3 |
| EOS% | 606 | 0.0 to 56.4 | 0.983 | 1.0 (0.976, 1.0) | 0.2 (0.2, 0.213) | 0.2 | 13.5 |
| EOS# x10<sup>3</sup>/μL | 606 | 0.0 to 34.55 | 0.999 | 1.0 (1.0, 1.045) | 0.02 (0.01, 0.02) | 0.02 | 16.0 |
| BASO% | 644 | 0.0 to 5.1 | 0.629 | 1.333 (1.2, 1.5) | -0.167 (-0.25, -0.14) | 0 | -8.1 |
| BASO# x10<sup>3</sup>/μL | 644 | 0.0 to 1.36 | 0.713 | 1.25 (1.043, 1.333) | -0.01 (-0.013, -0.001) | 0 | -0.1 |

BASO, basophil; CI, confidence interval; EOS, eosinophil; HCT, haematocrit; HGB, haemoglobin; LYMPH,
lymphocyte; MCHC, mean corpuscular haemoglobin concentration; MCH, mean corpuscular haemoglobin; MCV, mean corpuscular volume; MONO, monocyte; NEUT, neutrophil; PLT, platelet; WBC, white blood cell; RBC, red blood cell; RDW, red blood cell distribution width.

**Supplemental Table S3:** Sensitivity and specificity for WBC morphological abnormalities (blasts, immature granulocytes, nucleated RBCs, and atypical lymphocytes) compared to light microscopy.

|  | Flagged by OLO | Not flagged by OLO |  | % | 95% CI |  |
| --- | --- | --- | --- | --- | --- | --- |
| Positive by light microscopy | 93 | 7 | Sensitivity (PPA) | 93.0% | 86.1% | 97.1% |
| Negative by light microscopy | 21 | 87 | Specificity (NPA) | 80.6% | 71.8% | 87.5% |
|  |  |  | Overall Agreement | 86.5% | 81.1% | 90.9% |

**Supplemental Table S4:** Results of the reproducibility study, in which three levels of commercial control material (low, normal and high) were analyzed for all reported parameters. 240 samples were included for each level of control in these studies.

| Measurand | Level | Mean | Within run |  | Between Run |  | Between Day |  | Between instrument |  | Between Site |  | Total |  |
| --- | --- | --- | --- | --- | --- | --- | --- | --- | --- | --- | --- | --- | --- | --- |
|  |  |  | SD | CV% | SD | CV% | SD | CV % | SD | CV % | SD | CV% | SD | CV% |
| WBC | Low | 3.33 | 0.17 | 5.2 | 0.06 | 1.7 | 0.00 | 0.0 | 0.11 | 3.3 | 0.00 | 0.0 | 0.21 | 6.4 |
|  | Normal | 8.19 | 0.26 | 3.2 | 0.09 | 1.1 | 0.00 | 0.0 | 0.17 | 2.1 | 0.00 | 0.0 | 0.32 | 4.0 |
|  | High | 23.02 | 0.62 | 2.7 | 0.25 | 1.1 | 0.10 | 0.4 | 0.39 | 1.7 | 0.00 | 0.0 | 0.78 | 3.4 |
| RBC | Low | 2.61 | 0.04 | 1.5 | 0.04 | 1.7 | 0.00 | 0.0 | 0.07 | 2.8 | 0.00 | 0.0 | 0.09 | 3.6 |
|  | Normal | 5.14 | 0.05 | 0.9 | 0.04 | 0.8 | 0.00 | 0.0 | 0.10 | 1.9 | 0.00 | 0.0 | 0.12 | 2.2 |
|  | High | 5.91 | 0.06 | 1.0 | 0.03 | 0.6 | 0.02 | 0.4 | 0.10 | 1.7 | 0.00 | 0.0 | 0.12 | 2.1 |
| PLT | Low | 70.7 | 5.1 | 7.2 | 1.0 | 1.4 | 0.0 | 0.0 | 1.5 | 2.1 | 3.1 | 4.4 | 6.2 | 8.8 |
|  | Normal | 218.2 | 9.4 | 4.3 | 2.5 | 1.2 | 2.1 | 0.9 | 3.1 | 1.4 | 2.7 | 1.2 | 10.8 | 4.9 |
|  | High | 432.6 | 19.7 | 4.5 | 7.0 | 1.6 | 6.9 | 1.6 | 10.9 | 2.5 | 0.0 | 0.0 | 24.5 | 5.7 |
| HGB | Low | 7.05 | 0.10 | 1.4 | 0.09 | 1.2 | 0.00 | 0.0 | 0.10 | 1.4 | 0.00 | 0.0 | 0.16 | 2.3 |
|  | Normal | 14.41 | 0.10 | 0.7 | 0.05 | 0.4 | 0.02 | 0.1 | 0.11 | 0.7 | 0.11 | 0.8 | 0.19 | 1.3 |
|  | High | 17.32 | 0.13 | 0.7 | 0.06 | 0.3 | 0.05 | 0.3 | 0.11 | 0.6 | 0.20 | 1.2 | 0.27 | 1.6 |
| HCT | Low | 21.15 | 0.39 | 1.9 | 0.25 | 1.2 | 0.10 | 0.5 | 0.51 | 2.4 | 0.00 | 0.0 | 0.70 | 3.3 |
|  | Normal | 43.99 | 0.60 | 1.4 | 0.35 | 0.8 | 0.00 | 0.0 | 0.81 | 1.8 | 0.00 | 0.0 | 1.06 | 2.4 |
|  | High | 56.55 | 0.84 | 1.5 | 0.44 | 0.8 | 0.17 | 0.3 | 0.82 | 1.5 | 0.00 | 0.0 | 1.26 | 2.2 |
| MCV | Low | 80.91 | 0.84 | 1.0 | 0.74 | 0.9 | 0.00 | 0.0 | 0.79 | 1.0 | 0.00 | 0.0 | 1.37 | 1.7 |
|  | Normal | 85.62 | 0.81 | 1.0 | 0.72 | 0.8 | 0.00 | 0.0 | 0.98 | 1.1 | 0.00 | 0.0 | 1.46 | 1.7 |
|  | High | 95.66 | 0.86 | 0.9 | 0.52 | 0.5 | 0.00 | 0.0 | 1.51 | 1.6 | 0.00 | 0.0 | 1.82 | 1.9 |
| RDW | Low | 14.20 | 0.26 | 1.8 | 0.07 | 0.5 | 0.01 | 0.1 | 0.03 | 0.2 | 0.16 | 1.2 | 0.32 | 2.2 |
|  | Normal | 13.17 | 0.19 | 1.5 | 0.00 | 0.0 | 0.01 | 0.1 | 0.06 | 0.5 | 0.07 | 0.6 | 0.22 | 1.6 |
|  | High | 12.57 | 0.19 | 1.5 | 0.00 | 0.0 | 0.00 | 0.0 | 0.07 | 0.6 | 0.07 | 0.5 | 0.21 | 1.7 |
| MCH | Low | 26.99 | 0.19 | 0.7 | 0.17 | 0.6 | 0.05 | 0.2 | 0.49 | 1.8 | 0.00 | 0.0 | 0.55 | 2.1 |
|  | Normal | 28.04 | 0.18 | 0.6 | 0.15 | 0.5 | 0.00 | 0.0 | 0.44 | 1.6 | 0.00 | 0.0 | 0.50 | 1.8 |
|  | High | 29.30 | 0.22 | 0.7 | 0.04 | 0.1 | 0.05 | 0.2 | 0.46 | 1.6 | 0.00 | 0.0 | 0.52 | 1.8 |
| MCHC | Low | 33.37 | 0.38 | 1.1 | 0.26 | 0.8 | 0.00 | 0.0 | 0.60 | 1.8 | 0.00 | 0.0 | 0.76 | 2.3 |
|  | Normal | 32.76 | 0.39 | 1.2 | 0.23 | 0.7 | 0.02 | 0.1 | 0.46 | 1.4 | 0.00 | 0.0 | 0.65 | 2.0 |
|  | High | 30.63 | 0.40 | 1.3 | 0.15 | 0.5 | 0.00 | 0.0 | 0.32 | 1.0 | 0.31 | 1.0 | 0.62 | 2.0 |
| NEUT% | Low | 50.32 | 2.44 | 4.9 | 0.00 | 0.0 | 0.46 | 0.9 | 0.43 | 0.9 | 0.97 | 1.9 | 2.70 | 5.4 |
|  | Normal | 41.73 | 1.62 | 3.9 | 0.00 | 0.0 | 0.00 | 0.0 | 0.21 | 0.5 | 0.00 | 0.0 | 1.64 | 3.9 |
|  | High | 54.35 | 1.05 | 1.9 | 0.16 | 0.3 | 0.19 | 0.4 | 0.48 | 0.9 | 0.63 | 1.2 | 1.34 | 2.5 |
| NEUT# | Low | 1.68 | 0.12 | 7.0 | 0.02 | 1.4 | 0.03 | 1.7 | 0.05 | 3.2 | 0.00 | 0.0 | 0.13 | 8.0 |
|  | Normal | 3.42 | 0.16 | 4.7 | 0.05 | 1.3 | 0.00 | 0.0 | 0.06 | 1.9 | 0.00 | 0.0 | 0.18 | 5.2 |
|  | High | 12.51 | 0.39 | 3.1 | 0.14 | 1.2 | 0.10 | 0.8 | 0.26 | 2.1 | 0.16 | 1.3 | 0.53 | 4.2 |
| LYMPH% | Low | 17.62 | 1.69 | 9.6 | 0.29 | 1.6 | 0.00 | 0.0 | 0.00 | 0.0 | 0.00 | 0.0 | 1.71 | 9.7 |
|  | Normal | 36.41 | 1.42 | 3.9 | 0.00 | 0.0 | 0.30 | 0.8 | 0.00 | 0.0 | 0.29 | 0.8 | 1.48 | 4.1 |
|  | High | 25.30 | 0.91 | 3.6 | 0.00 | 0.0 | 0.17 | 0.7 | 0.07 | 0.3 | 0.00 | 0.0 | 0.93 | 3.7 |
| LYMPH# | Low | 0.59 | 0.06 | 10.4 | 0.02 | 3.7 | 0.00 | 0.0 | 0.01 | 2.5 | 0.00 | 0.0 | 0.07 | 11.4 |
|  | Normal | 2.98 | 0.15 | 5.2 | 0.01 | 0.5 | 0.02 | 0.7 | 0.08 | 2.5 | 0.00 | 0.0 | 0.17 | 5.8 |
|  | High | 5.82 | 0.28 | 4.7 | 0.00 | 0.0 | 0.03 | 0.6 | 0.09 | 1.5 | 0.00 | 0.0 | 0.29 | 5.0 |
| MONO% | Low | 12.21 | 1.53 | 12.5 | 0.00 | 0.0 | 0.22 | 1.8 | 0.73 | 6.0 | 0.59 | 4.9 | 1.81 | 14.8 |
|  | Normal | 8.44 | 0.89 | 10.6 | 0.14 | 1.6 | 0.07 | 0.9 | 0.30 | 3.6 | 0.43 | 5.1 | 1.05 | 12.4 |
|  | High | 11.88 | 0.76 | 6.4 | 0.00 | 0.0 | 0.00 | 0.0 | 0.42 | 3.6 | 0.72 | 6.0 | 1.12 | 9.5 |
| MONO# | Low | 0.41 | 0.05 | 13.4 | 0.00 | 0.0 | 0.00 | 0.0 | 0.04 | 9.3 | 0.01 | 1.5 | 0.07 | 16.4 |
|  | Normal | 0.69 | 0.08 | 11.2 | 0.02 | 2.3 | 0.00 | 0.0 | 0.04 | 5.2 | 0.03 | 3.7 | 0.09 | 13.1 |
|  | High | 2.73 | 0.19 | 7.0 | 0.03 | 0.9 | 0.00 | 0.0 | 0.12 | 4.4 | 0.14 | 5.0 | 0.26 | 9.7 |
| EOS% | Low | 11.75 | 1.69 | 14.4 | 0.30 | 2.6 | 0.00 | 0.0 | 0.00 | 0.0 | 0.41 | 3.5 | 1.76 | 15.0 |
|  | Normal | 7.09 | 0.85 | 11.9 | 0.19 | 2.7 | 0.00 | 0.0 | 0.08 | 1.1 | 0.14 | 2.0 | 0.88 | 12.5 |
|  | High | 3.94 | 0.47 | 11.9 | 0.00 | 0.0 | 0.00 | 0.0 | 0.00 | 0.0 | 0.00 | 0.0 | 0.47 | 11.9 |
| EOS# | Low | 0.39 | 0.06 | 16.2 | 0.01 | 2.2 | 0.00 | 0.0 | 0.02 | 5.3 | 0.01 | 1.3 | 0.07 | 17.2 |
|  | Normal | 0.58 | 0.07 | 12.4 | 0.02 | 3.9 | 0.00 | 0.0 | 0.02 | 2.8 | 0.00 | 0.0 | 0.08 | 13.3 |
|  | High | 0.91 | 0.11 | 12.6 | 0.00 | 0.0 | 0.01 | 1.4 | 0.01 | 0.6 | 0.00 | 0.0 | 0.12 | 12.7 |
| BASO% | Low | 8.12 | 1.23 | 15.1 | 0.00 | 0.0 | 0.20 | 2.5 | 0.03 | 0.3 | 0.21 | 2.6 | 1.26 | 15.5 |
|  | Normal | 6.33 | 0.75 | 11.9 | 0.00 | 0.0 | 0.00 | 0.0 | 0.00 | 0.0 | 0.16 | 2.5 | 0.77 | 12.1 |
|  | High | 4.54 | 0.48 | 10.5 | 0.13 | 2.8 | 0.00 | 0.0 | 0.00 | 0.0 | 0.12 | 2.5 | 0.51 | 11.2 |
| BASO# | Low | 0.27 | 0.04 | 15.4 | 0.01 | 2.3 | 0.01 | 1.9 | 0.00 | 1.8 | 0.00 | 1.0 | 0.04 | 15.8 |
|  | Normal | 0.52 | 0.07 | 12.6 | 0.00 | 0.0 | 0.00 | 0.0 | 0.01 | 1.7 | 0.01 | 2.6 | 0.07 | 12.9 |
|  | High | 1.04 | 0.11 | 10.7 | 0.04 | 3.5 | 0.00 | 0.0 | 0.03 | 3.1 | 0.02 | 2.4 | 0.12 | 11.9 |

BASO, basophil; CI, confidence interval; EOS, eosinophil; HCT, haematocrit; HGB, haemoglobin; LYMPH, lymphocyte; MCHC, mean corpuscular haemoglobin concentration; MONO, Monocyte; MCH, mean corpuscular haemoglobin; MCV, mean corpuscular volume; NEUT, neutrophil; PLT, platelet; RBC, red blood cell; RDW, red blood cell distribution width; WBC, white blood cell.

## Notes

### Clinical Trial

NCT03595501

### Author Declarations

Boston Children's Hospital Institutional Review Board, Boston, MA, USA Presbyterian Healthcare Services Human Research Protections and Institutional Review Board, Albuquerque, NM, USA Columbia University Human Research Protection Office Institutional Review Boards, New York, NY, USA Tel Aviv Sourasky Medical Center Ethics Comittee, Tel Aviv, Israel

